# Pulmonary Radioaerosol Mucociliary Clearance Parameters as Potential Outcomes in Primary Ciliary Dyskinesia Trials

**DOI:** 10.1101/2023.06.05.23290928

**Authors:** June K. Marthin, Mathias G. Holgersen, Kim G. Nielsen, Jann Mortensen

## Abstract

**Background:** Pulmonary Radioaerosol Mucociliary Clearance (PRMC) is an *in vivo* whole lung ciliary function test reliable for assessing mucociliary clearance for diagnostic purposes in individuals suspected of primary ciliary dyskinesia (PCD). We aimed to evaluate expanded use of PRMC by providing advantages and limitations for its potential use in providing outcome parameters in future trials aiming to restore ciliary activity.

**Material and Methods:** In this retrospective study, PRMC tests performed over a period of 24 years (1999-2022) were meticulously re-analyzed. Patients with genetically verified PCD and non-PCD controls were included. Originally, nebulized ^99m^Tc-albumin colloid was inhaled, and static and dynamic imaging acquired for 60 and 120 minutes, and 24 hours.

For the purpose of the present study 3 PRMC parameters were defined: 1 hour lung retention (LR1), tracheobronchial velocity (TBV), and cough clearance.

**Results:** Sixty-nine patients were included from the Danish PCD cohort. PRMC was overall completely absent regardless of PCD genotypes. In one patient with CCDC103 mutation, residual ciliary function and normal nasal NO, we found normal PRMC LR1 and measurable, however low, TBV.

Voluntary cough significantly increased clearance with a median (IQR) of 11 (4;24) %.

**Conclusion:** Absolute absence of PRMC would be the expected baseline result in by far the majority of patients with PCD regardless of genotype before introducing ciliary protein correctors in a clinical trial.

Measurable PRMC TBV and normal LR1 in one patient with residual ciliary function, indicated that PRMC parameters could potentially improve if ciliary function was to be restored during a clinical trial.

Involuntary cough and peripheral radioaerosol deposition were the main challenges of the PRMC method.

## INTRODUCTION

Primary ciliary dyskinesia (PCD; MIM244400) is a multisystem ciliopathy that includes dyskinetic respiratory cilia. The resulting impairment of mucociliary clearance leads to chronic destructive infectious and inflammatory disease of the upper and lower airways. To date, approximately 50 genes have been identified to harbor variants attributed to cause PCD (1-3). Most mutations associated with PCD are autosomal recessive. Less frequently, X-chromosomal recessive and *de novo* autosomal dominant inheritance has been described (4).

For more than two decades, the PRMC technique has been used as a supplementary diagnostic tool in the investigation of PCD in the national Danish PCD center. In patients with PCD (pwPCD), PRMC has effectively demonstrated abnormal clearance patterns resulting from impaired mucociliary clearance caused by abnormal ciliary motility. The technique is non-invasive and involves minimal radiation exposure of less than 1 mSv and visualizes whole-lung retention of the radioaerosol by use of a gamma-camera. In previous studies PRMC has shown high sensitivity to predict PCD and as a strong tool to rule out PCD in non-PCD cases, applicable to children as young as 5 years of age (5-9).

Given that pharmaceutical biotech companies are showing increasing interest in developing ciliary protein correctors for PCD, the need for relevant outcome parameters in future clinical trials has become increasingly apparent. Currently, PRMC is the only available method for assessing whole lung mucociliary clearance (10) and serves as an *in vivo* marker of the universal ciliary function throughout the entire compartment of ciliated airways. While PRMC has been used in randomized clinical trials in cystic fibrosis (11-12), it also holds potential as an outcome parameter for future clinical trials in PCD. Long term lung function decline in pwPCD is known to be heterogeneous (13-14) and growing evidence indicates that specific PCD gene defects may be related to lung disease outcomes (14-16). With patient specific therapies targeting specific PCD genes on the horizon, one of the major tasks in PCD research currently and in the years to come will be to identify genotype-phenotype relationships within subgroups of pwPCD. Variations in PRMC could potentially uncover specific genotype-phenotype relations enabling to explain differences in the expression of PCD disease.

## HYPOTHEZIS AND AIMS

We hypothesized that different PCD genotypes may result in measurable differences in mucociliary clearance, and thus, our primary aim was to investigate PRMC in patients with genetically confirmed PCD from the Danish PCD Cohort, and at the same time describe advantages and limitations of the PRMC method in delivering potential outcome parameters for future clinical PCD trials.

Our secondary aim was to investigate the impact of cough on PRMC measures and thereby also the efficacy of pulmonary mucociliary cough clearance in pwPCD. While pwPCD are often encouraged to cough up mucus as a compensatory mechanism for their impaired ciliary transport, the extent to which this may improve clearance has so far not been thoroughly studied.

## MATERIAL AND METHODS

### Study design

This was a retrospective, cross-sectional study conducted at a single center utilizing data from the Danish PCD Registry. The study involved the re-analyzis of historic diagnostic PRMC tests performed over a period of 24 years (1999-2022). Additional exploratory diagnostic test results such as PCD genetics and nasal Nitric Oxide (nNO) obtained during outpatient visits throughout the elapsed period were added to the data set.

Inclusion criteria:

*Patients with PCD above five years of age were eligible when fulfilling following criteria:*

1. Genetically confirmed bi-allelic mutations in a known PCD-causing gene.
2. A previous PRMC test conducted as part of PCD work-up due to a clinical suspicion of PCD providing that one or more of the defined PRMC outcome parameters: 1 hour lung retention (LR1), tracheobronchial velocity (TBV), or cough clearance could be determined with good quality.
3. Clinical symptoms consistent with PCD as well as available nNO, High Speed Video Microscopy (HSVM) and Transmission Electron Microscopy (TEM) diagnostic tests compatible with PCD in accordance with the current ERS criteria for diagnosis of PCD (17).

*Non-PCD controls:*

For the non-PCD control group, all PRMC tests conducted in the year 2022 were retrieved. Patients who underwent PRMC testing performed in this period as part of their PCD work-up but where PCD was finally ruled out, including lack of any mutations in a known PCD gene, were included in the control group as non-PCD referrals. An additional prerequisite was a conclusive PRMC test showing normal LR1 and TBV, and normal 24-hour lung retention.

### PRMC test: delivered outcome parameters

PRMC tests performed using identical technique and procedure throughout the 24 years of data acquisition were re-analyzed with a view to assess the following three defined parameters: PRMC LR1, PRMC TBV, and PRMC voluntary controlled and involuntary random cough clearance.

Whereas PRMC LR1 and TBV measurements were confined to only 60 minutes, cough clearance investigation was prolonged to 120 minutes.

#### PRMC LR1 assessment

The PRMC imaging technique has previously been described (5,7,10). In brief, patients inhaled ultrasonically (model 35 B; Devilbiss; Somerset, PA) nebulized ^99m^Tc-albumin colloid (Venticoll; GI PHARMA; Saluggia, Italy) during tidal breathing by 20 slow inspirations and forced expirations. Immediately following this radioaerosol inhalation procedure, subjects were placed in supine position against a posteriorly positioned gamma camera for detecting lung radioactivity. A complete measurement of pulmonary radioactivity was then performed as dynamic and static acquisitions within the first hour, the static acquisitions performed at 0, 30 and, 60 minutes. PRMC was then calculated over both lungs as whole-lung retention after 1 hour (LR1) corrected for background and physical decay. Cough was strictly monitored by trained staff within these hours. Patients were not allowed to leave the room and every cough was precisely documented as per minutes after inhalation of the initial tracer. This allowed ruling out periods (minutes) of coughing in the following assessment of the dynamic acquisitions, which was crucial to avoid false positive clearance results due to cough clearance.

From doorstep to doorstep a PRMC test lasted two hours: this included welcoming the patient, placing reference (^57^Co) markers on neck and back of the patient and taking the “zero”-acquisition with the markers, training the patient to do correct slow inhalation, and forced exhalation technique with isotonic saline, inhale the ^99m^Tc-albumin colloid radioaerosol, measure static and dynamic PRMC acquisitions, monitor for cough, removing the reference markers and sending the patient home.

Figure 1 shows examples of tracer movement from static acquisitions at 0, 30, and 60 minutes after inhalation in a patient with PCD and a non-PCD referral.

**Figure 1.**
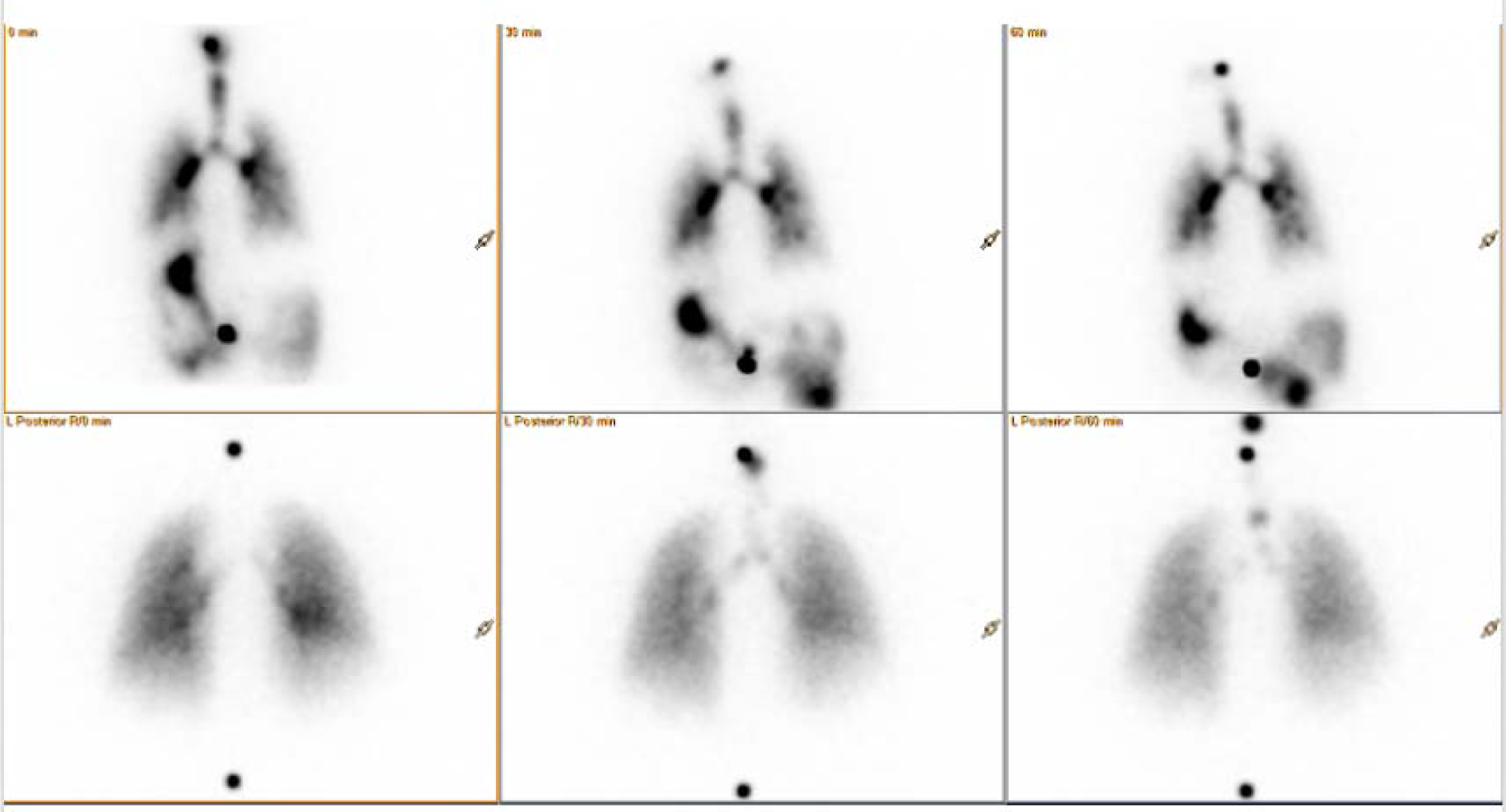
A series of static acquisitions showing radioaerosol tracer movement due to mucociliary transport in a patient with PCD (top row) and in a non-PCD individual (bottom row). *Top row*: A patient with PCD. The radioaerosol is seen initially centrally deposited (0 min), and still, there is almost no movement of the tracer after 30 minutes (middle) and 60 minutes (right), indicating that mucociliary clearance was minimal. The swallowed radioaerosol is seen to pass from the left-sided stomach further to the duodenum and jejunum. Images are taken posteriorly. *Bottom row*: A non-PCD referral where PCD was later ruled out. Centrally deposited radioaerosol inhaled at 0 minutes is quickly removed from the central airways, ascending upwards from the main bronchi to the trachea from where it is further cleared. The initial tracer foci after inhalation are markedly cleared already after 30 minutes (middle) and further cleared after 60 minutes (right) with only little radioaerosol left in the central airways, although some foci are still visible in the main bronchi and the trachea. *Top row and bottom row*: Two ^57^Co markers seen over the cervical and lumbar spine were placed initially to help positioning the subjects.

Regional ventilation distribution was visualized by a ^81m^Kr-gas scintigram. The initial ^99m^Tc aerosol distribution was compared to the ^81m^Kr ventilation distribution to indicate how far the radioaerosol had penetrated the airways and lungs and hence had to move before being cleared from the airways. This penetration index (PI) was used to calculate each subject’s predicted values for lung retention from our previously published reference equations (10) (Appendix 1).

In case of repeated PRMC measurements in the same patient, the measurement with the most central tracer distribution specified by the PI, was chosen.

#### Determination of PRMC TBV

To determine PRMC tracheobronchial velocity (TBV) we identified distinctive radiomucous boli formed in the tracheobronchial area that further ascended within the main bronchi and the trachea (OLS Supplemental file: movie 1a in a PCD patient and 1b in a non-PCD referral) which allowed for determination of PRMC TBV, if present. Two dynamic acquisitions of 20 minutes duration obtained within the first hour of the PRMC investigation were reviewed and analyzed for all included patients. Two readers re-analyzed each acquisition in a blinded fashion to the original diagnostic report. The transport distance between each frame was measured for any upward boli transport in the main bronchi and trachea.

As 1 frame equals 2 minutes (10 frames in a 20-minute acquisition), the PRMC TBV was calculated in mm/minutes using the following equation:

PRMC TBV = mm of bolus transportation / (Y frames * 2 minutes).

#### PRMC voluntary controlled and involuntary random cough clearance

Voluntary controlled cough clearance was performed in a subset of patients as repeatable cough maneuvers at the end of the PRMC measurements.

To avoid false negative results caused by cough clearance, any cases of involuntary random cough during the first hour of a PRMC test were discarded.

We compared these previously discarded PRMC tests for lung retention values at 30, 60 and 120 minutes with PRMC measurements in patients who did not cough during the PRMC investigation. This allowed us to analyze the impact of involuntary cough on PRMC.

### Additional diagnostic testing

#### PCD Genetics

The following PCD genes were analysed:

ARMC4, C21orf59, CCDC103, CCDC114, CCDC151, CCDC65, CCDC39, CCDC40, CCNO, CENPF, DNAH11, DNAH5, DNAH9, DNAI1, DNAI2, DNAL1, DNAAF1, DNAAF2, DNAAF3, DNAAF4, DRC1, GAS8, HYDIN, INVS, LRRC6, MCIDAS, OFD1, PIH1D3, RSPH1, RSPH3, RSPH4A, RSPH9, SPAG1, ZMYND10, and FOXJ1.

#### Nasal NO measurement

nNO was measured using the stationary NO chemiluminescence analyzer CLD88sp (ECO MEDICS® AG, Duernten, Switzerland) and according to the recommended technical standards (18).

nNO was measured in aspirated air via a nasal olive probe inserted into one nostril. Mean nNO concentration in parts per billion (ppb) was calculated from triple measurements in each subject. All subjects had nNO sampled during velum closure, preferentially by exhalation against resistance, and if this was not possible for a subject, sampling was performed during breath hold.

The sampling flow rate was 0.33 L·min^−1^ and conversion from nNO concentration in ppb to nNO production rate (nL·min^−1^) was calculated as follows:

*nNO (ppb)×flowrate (L·min^−1^) =nNO (nL·min^−1^)*.

Ambient NO was recorded before each measurement.

#### High Speed Video Microscopy (HSVM) and Transmission Electron Microscopy (TEM)

HSVM ciliary function analysis and TEM was performed as standard diagnostic testing in all pwPCD included in this study and according to guidelines (17,19-20).

### Statistical analysis

Non-parametric statistics were used to analyse the data due to the small sample size. Fisher’s exact test was used to compare count data in tables, while the Kruskal-Wallis test was used to compare three or more independent groups. Wilcoxon rank-sum test was used to compare means of individual and independent groups.

All statistical analyses were performed using R version 4.2.0. P-values < 0.05 was considered statistically significant.

### Ethics

The study did not require approval from the ethical authorities since the only data used was existing retrospective registry data (informed consent from all patients) previously collected for diagnostic purpose in relation to clinical PCD work-up.

## RESULTS

We included 69 patients from our Danish PCD cohort who had previously undergone a PRMC test, out of a total of 82 patients with genetically confirmed PCD and a previous PRMC test. Among these 69 patients, we were able, based on judgements of quality, to include 49 for PRMC LR1 radioaerosol retention assessments, 34 for dynamic PRMC TBV assessments, and 17 for cough clearance investigation (Table 1 and Figure 2). Additionally, we included 6 non-PCD referrals as controls for comparison of PRMC TBV values.

**Figure 2.**
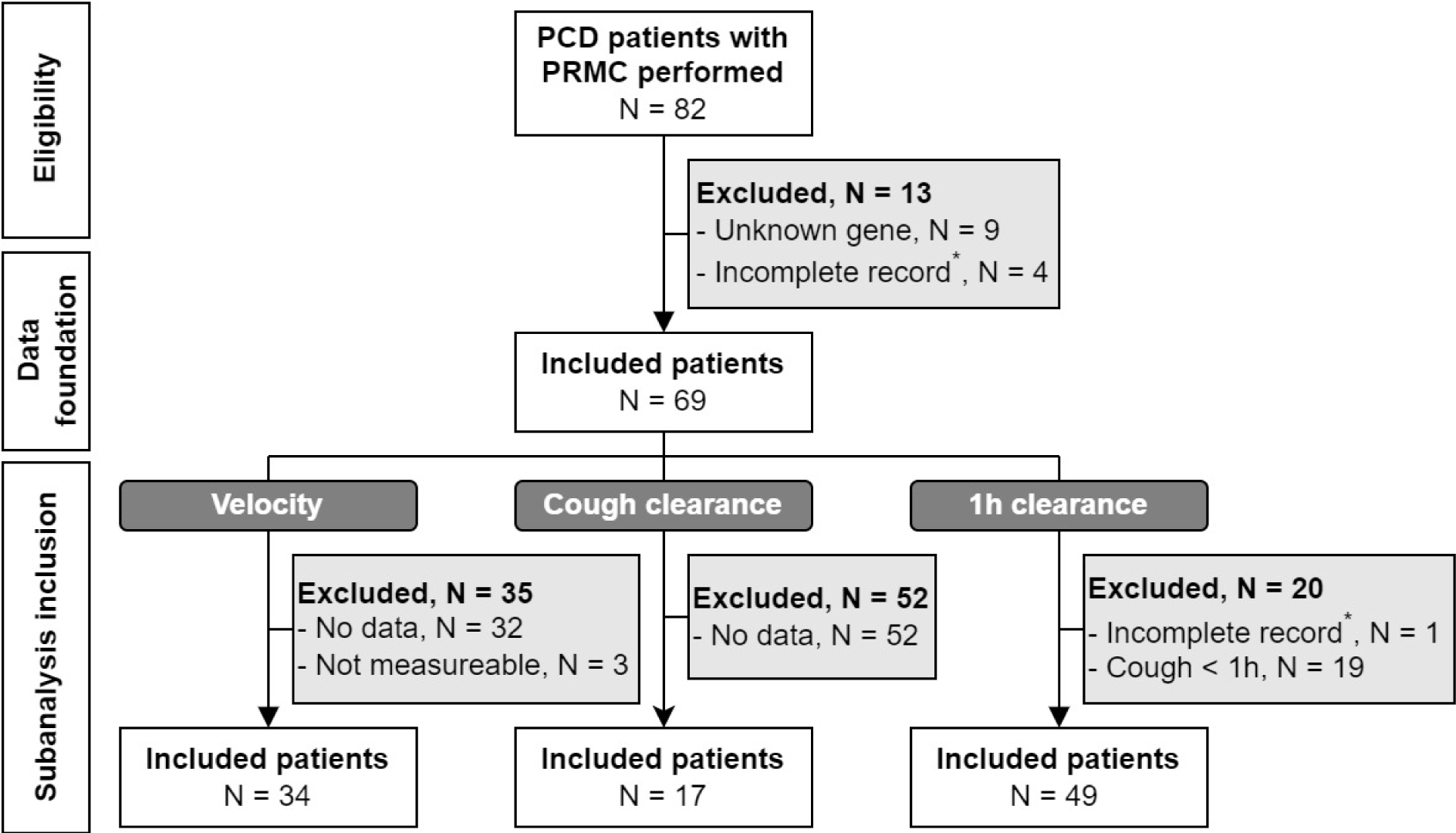
Flowchart of patient inclusion. A total of 82 patients with PCD had a historically performed Pulmonary Radioaerosol Mucociliary Clearance (PRMC) test. Thirteen patients were excluded, nine due to unknown genetics and four due to an incomplete cough record*. Thereby, a total of 69 patients constitutes the data in the study. Thirty-four patients were included for velocity measurement, 17 patients were included for voluntary cough clearance measurement and 49 patients were included for conclusive assessment of 1 hour lung retention (LR1).

**Table 1.**
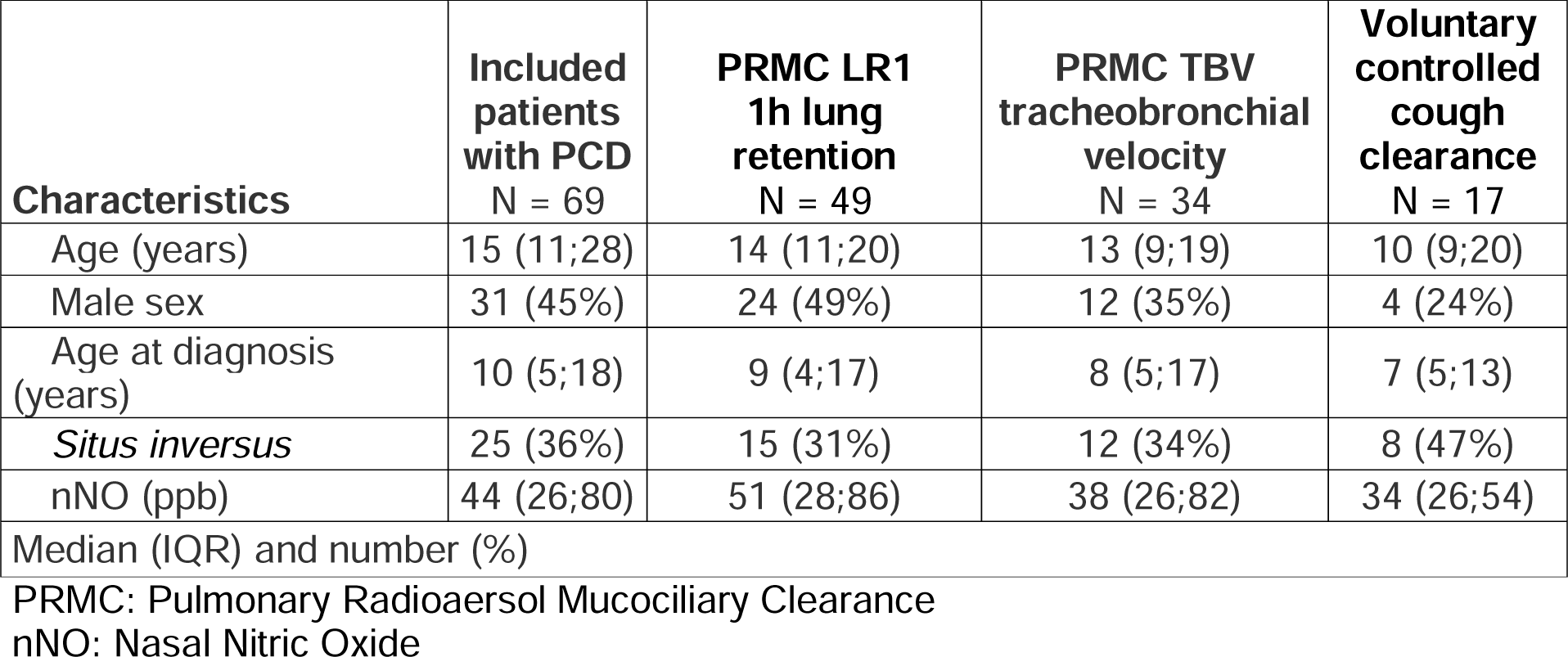
Characteristics and numbers of patients with PCD included in the overall study and each sub-study based on outcome parameter.

| **Characteristics** | **Included patients with PCD**<br>**N = 69** | **PRMC LR1**<br>**1h lung retention**<br>**N = 49** | **PRMC TBV**<br>**tracheobronchial velocity**<br>**N = 34** | **Voluntary controlled cough clearance**<br>**N = 17** |
| --- | --- | --- | --- | --- |
| Age (years) | 15 (11;28) | 14 (11;20) | 13 (9;19) | 10 (9;20) |
| Male sex | 31 (45%) | 24 (49%) | 12 (35%) | 4 (24%) |
| Age at diagnosis (years) | 10 (5;18) | 9 (4;17) | 8 (5;17) | 7 (5;13) |
| *Situs inversus* | 25 (36%) | 15 (31%) | 12 (34%) | 8 (47%) |
| nNO (ppb) | 44 (26;80) | 51 (28;86) | 38 (26;82) | 34 (26;54) |
| Median (IQR) and number (%) |  |  |  |  |

PRMC: Pulmonary Radioaerosol Mucociliary Clearance
nNO: Nasal Nitric Oxide

### PRMC 1 hour lung retention (LR1) assessment

Forty-nine out of 69 (71%) patients were able to complete the first hour of PRMC testing without coughing and were included in the analysis of PRMC 1 hour retention in relation to genotypes and proxy measures (HSVM ciliary motility patterns and TEM ultrastructure), Figure 3.

**Figure 3.**
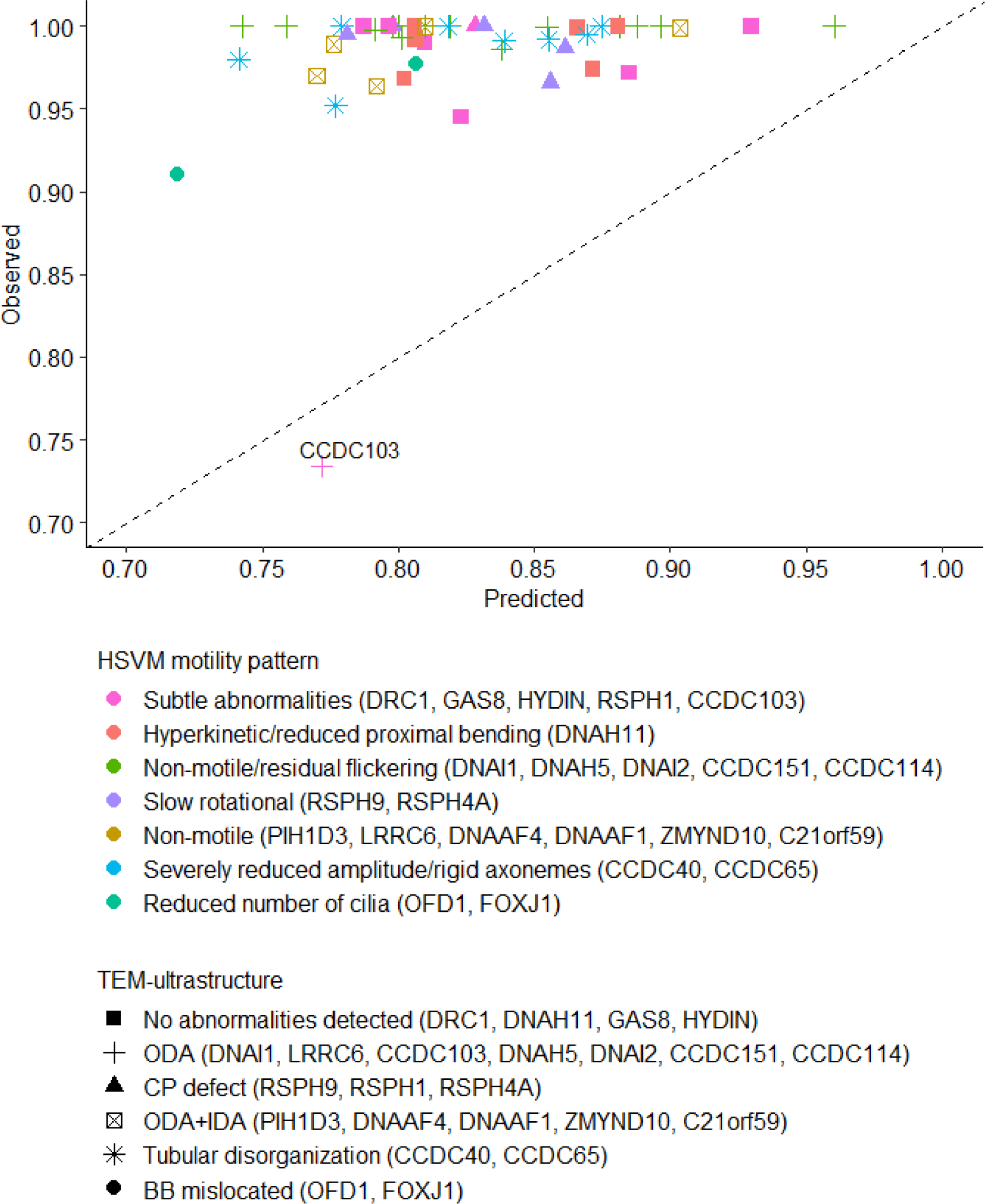
1-hour lung retention and indirect genotype measures. Measured 1 hour lung retention (LR1) across genotypes and TEM groups (y-axis) in relation to predicted normal values calculated based on aerosol deposition (penetration index), age and sex. The dotted line indicates alignment of measured and predicted values, meaning points are expected to lie on the line for healthy subjects. The colors indicate the observed ciliary beat patterns as assessed by high-speed video microscopy (HSVM). Shapes indicate the observed ciliary ultrastructure as assessed by transmission electron microscopy (TEM). In parenthesis, the genotypes are given.

We compared the five most prevalent genotypes CCDC39/CCDC40, DNAH11, DNAH5, DNAI1 and HYDIN, and found no significant differences either in the observed LR1 values or the retention z-scores (Table 2).

**Table 2.**
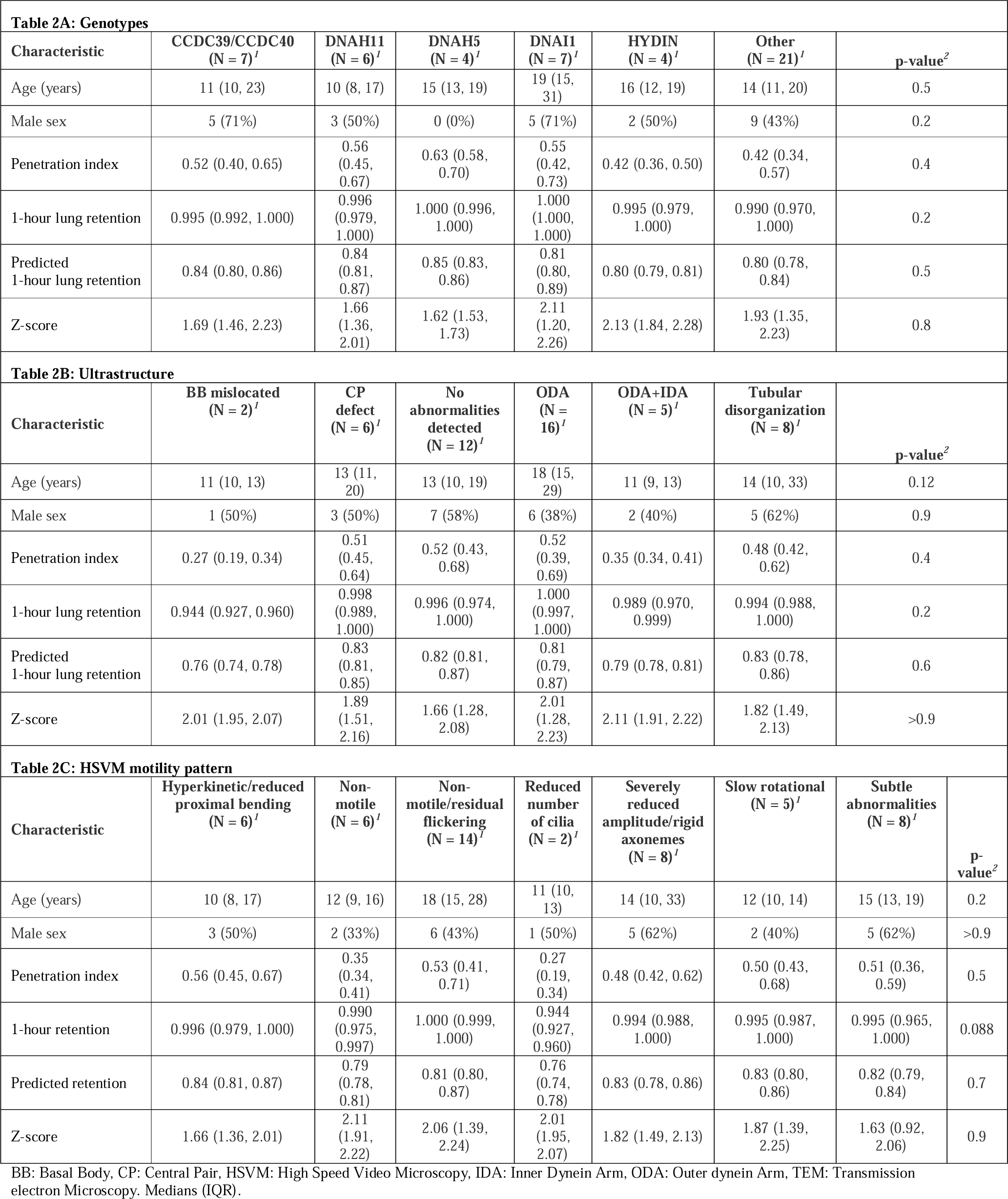
PRMC 1 hour retention (LR1) and association with genotypes, TEM and HSVM

Subgroup analyzes of PRMC values across TEM and HSVM groups also revealed no significant differences.

In one patient with CCDC103 defect LR1 was significantly lower as it reached normal value (Figure 3).

The complete list of in- and excluded patients according to genotypes, HSVM ciliary motility patterns and TEM ultrastructure is provided in Supplemental file Figure 2.

### PRMC tracheobronchial velocity

Thirty-four patients were included for dynamic PRMC assessment, of which only one patient with PCD had a measurable PRMC TBV of 1.5 mm/min, while the rest of the patients had no measurable velocity i.e., 0 mm/min, Figure 4.

**Figure 4:**
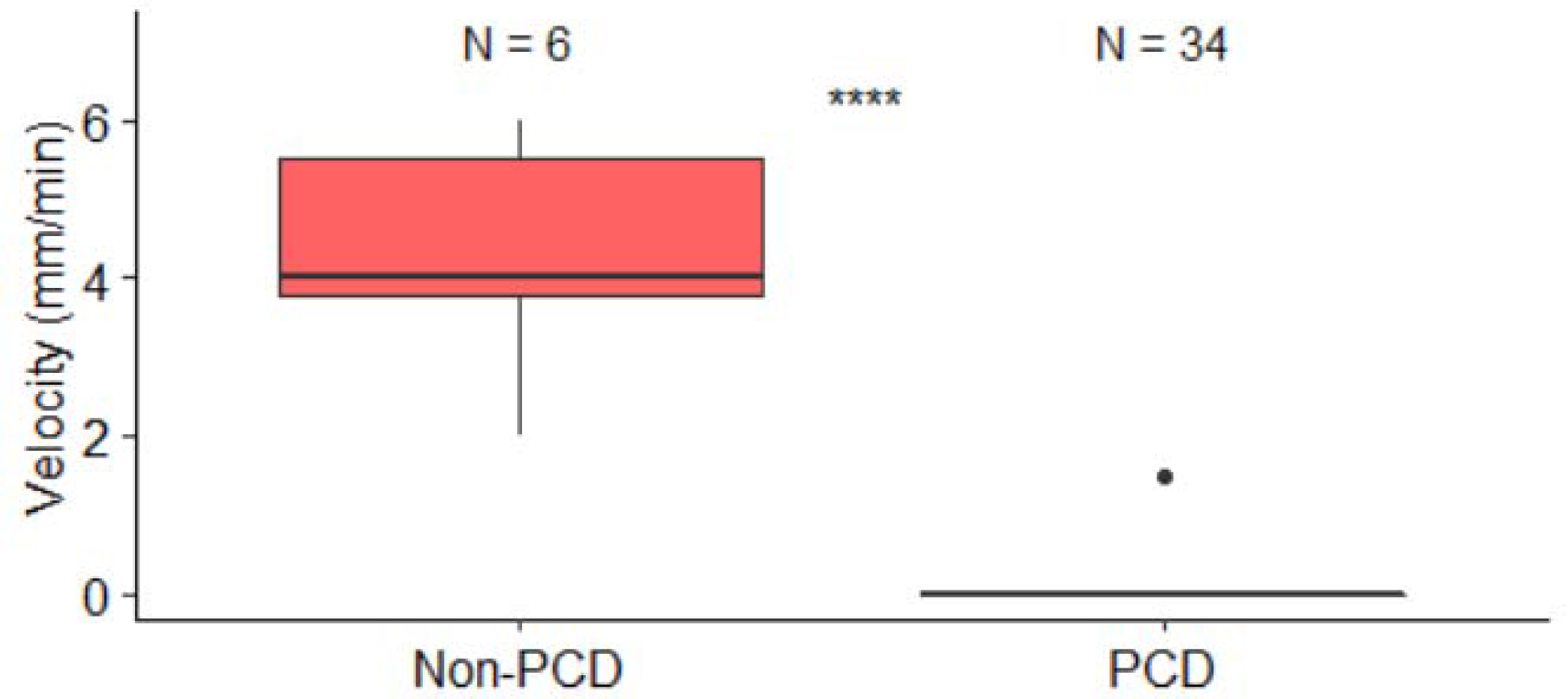
Comparison of PRMC velocity between included patients with PCD and non-PCD controls. PRMC tracheobronchial velocity (TBV) was 0 mm/min in all patients except one in PCD (33/34, 97%). The one patient with PCD and measurable PRMC TBV was homozygous for p.His154Pro mutation in the CCDC103 gene and had normal nasal NO of 967 ppb (319 nL/min). The PRMC TBV in this patient was significantly lower (1.5 mm/min) compared to non-PCD referral PRMC TBV (median 4.0 mm/min, range 2.0 to 6.0 mm/min), P< 0.00001.

This patient had a homozygous p.His154Pro CCDC103 defect, normal nNO levels of 968 ppb (319 nL/min), and residual (subtle abnormal) ciliary beating with a low-to-normal CBF of 3.5-8.9 Hz on HSVM, and a moderate ODA deficiency (TEM: mean ODA count of 5.3 (>7). The PRMC TBV of 1.5 mm/min in this patient was significantly lower compared to non-PCD referrals in the control group who had a median (range) PRMC velocity of 4.0 (2.0 to 6.0 mm/min, P<0.00001), Figure 4.

### PRMC Cough clearance

Voluntary controlled cough clearance from the central parts of the lungs was found to be median (IQR) 11 (4;24) % in 17 pwPCD, indicating the usefulness of deliberate cough as an advantageous mechanism for mucus clearance even in PCD. A statistically significant moderate negative linear correlation between central cough clearance and the PI was found, indicating that a centrally deposited aerosol (low PI value) is more easily coughed up from the main bronchi and the trachea than a peripherally deposited aerosol (high PI value), Figure 5.

**Figure 5:**
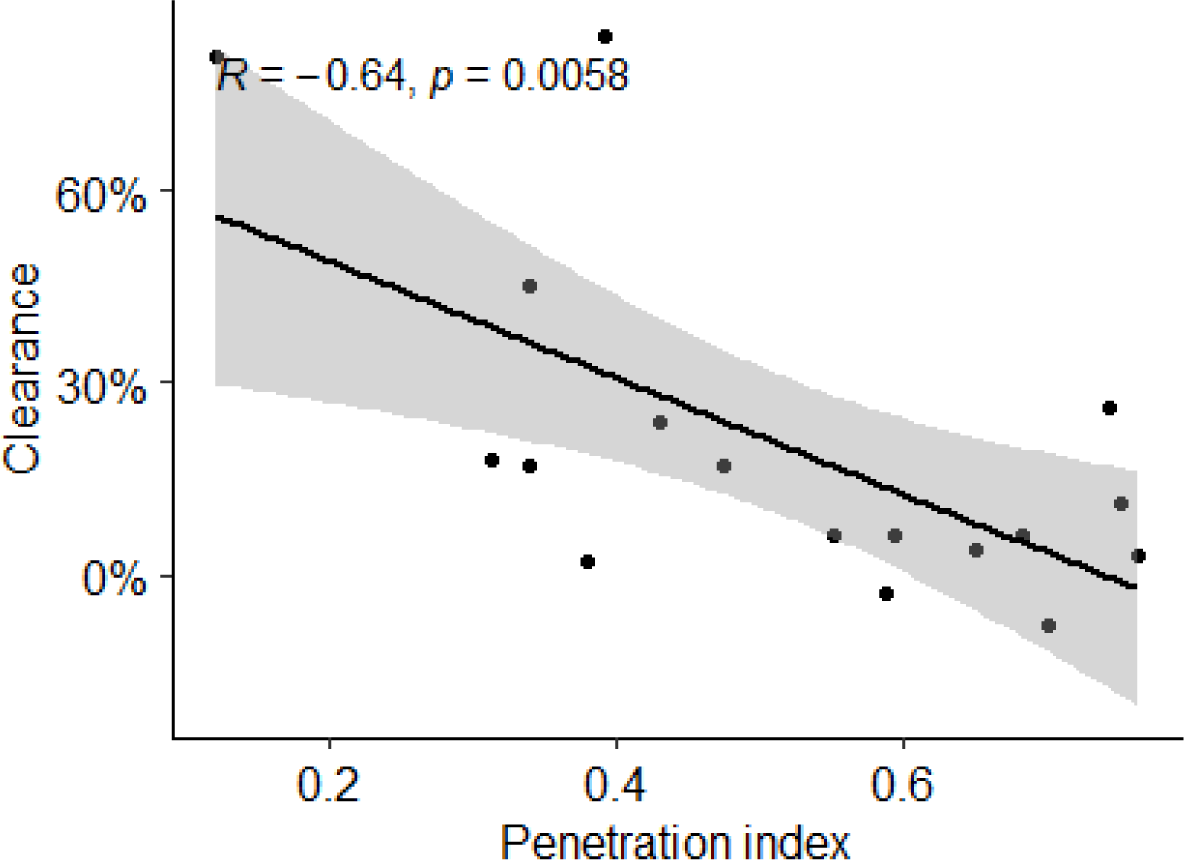
Correlation between voluntary cough-clearance and aerosol deposition. Cough clearance in relation to penetration index (PI) measured in 17 patients. As the penetration index increases (more peripheral radioaerosol distribution), the cough clearance decreases. The moderate negative correlation was significant (P = 0.0058). The strength of the relationship is represented by the slope of the solid line and denoted by the Pearson correlation coefficient.

### Effect of involuntary cough on PRMC

Also, involuntary random cough resulted in significant cough clearance. A comparison of patients who did not cough to those who coughed during PRMC showed a significant difference in measured lung retention at all timepoints, with cough clearance increasing over time, Figure 6.

**Figure 6:**
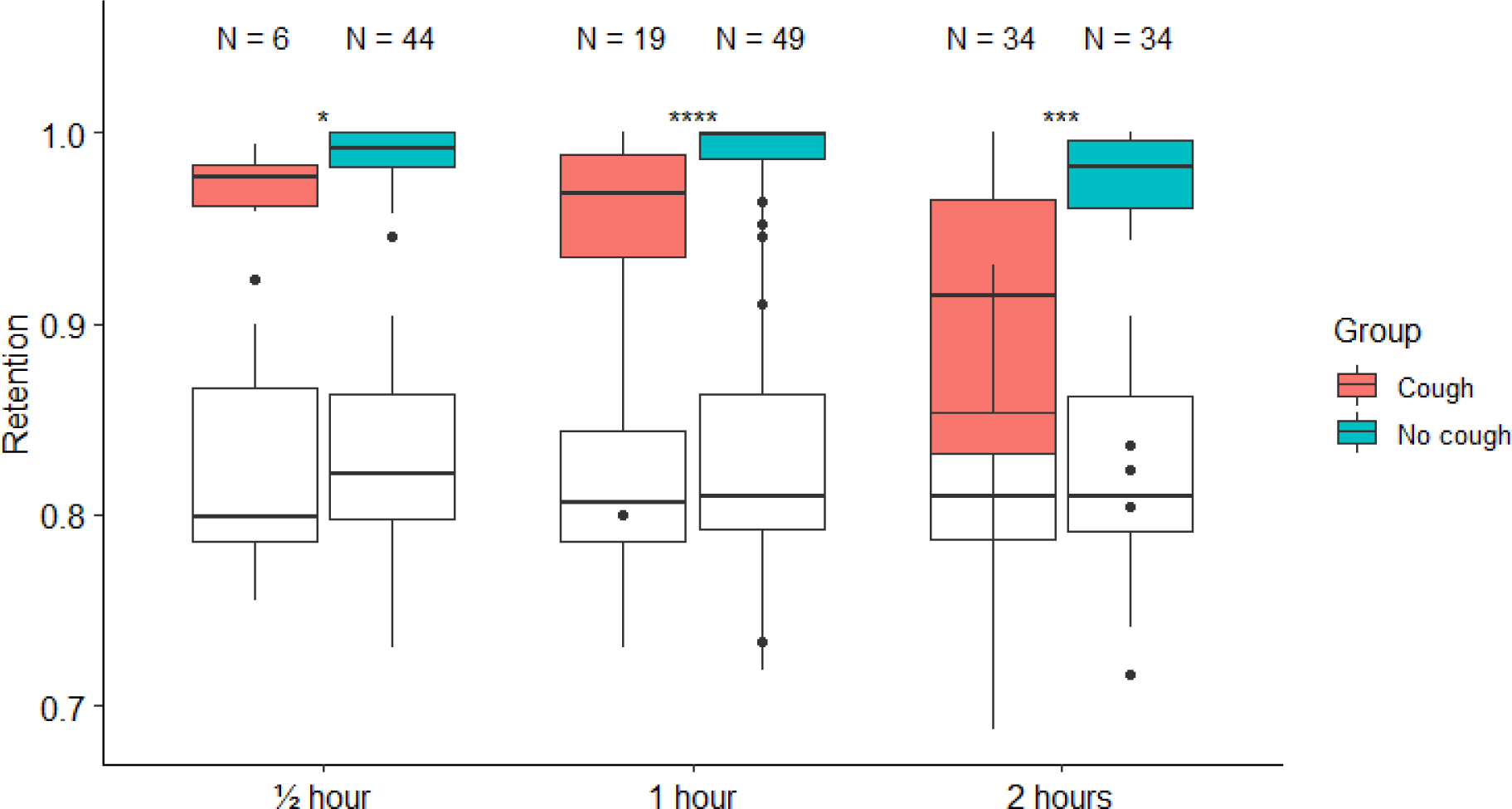
Boxplots of retention measurements for the included patients with PCD stratified by records of cough. Boxplots showing measured (colored boxplots) and predicted (white boxplots) lung retention at 3 timepoints (30 minutes, 1 hour and 2 hours) for patients with no records of coughing (blue boxes) and patients who did cough (red boxes) during PRMC measurements. The figure shows how lung retentions reach toward normal values (red boxes) in the patients with records of cough, indicating measurable and increasing lung clearance due to cough. Level of significance *;***;**** according to text.

The median lung retention values for patients with no recorded cough versus recorded cough were as follows: 30-minute LR: 99.2% (98.1-100) retention versus 97.6% (96.2-98.3) retention (P = 0.02544), 60-minute LR: 99.9% (98.6-100) retention versus 96.8% (93.5-98.8) retention (P < 0.0001), and 120-minute LR: 98.2% (96-99.6) retention versus 91.4% (83.2-96.5) retention (P = 0.0006). The differences in lung retention between the two groups could not be explained by differences in PI, which were similar between groups at all time points (at 30 min: P = 0.7653, at 60 min: P = 0.9778 and at 120 min: P= 0.8952).

## DISCUSSION

This study investigated PRMC in 69 patients with genetically confirmed PCD. We here present a description of various PRMC parameters across genotypes. From our experience with the PRMC method in PCD for more than 2 decades we also put into perspective the advantages and pitfalls of this method, with regards to a range of PRMC parameters potentially appropriate as study outcomes in future clinical trials in pwPCD. Overall, our results showed no significant differences in LR1 values or retention z-scores among the five most prevalent genotypes, nor when grouping patients based on TEM ultrastructure or ciliary motility patterns in HSVM. We found that coughing, an often unavoidable and difficult to suppress defense mechanism for pwPCD, not surprisingly significantly affected PRMC, with voluntary coughing resulting in a median clearance of 11%, while involuntary coughing resulted in significant clearance at all time points measured.

### PRMC geno-and phenotype

Overall, our study showed that PRMC was extremely low or unmeasurable in all parameters regardless of PCD genotype. Subgroup analysis for TEM groups and ciliary motility patterns by HSVM did not show differences across groups, either. This is interesting, since previous studies have shown that certain PCD genes can be associated with either more severe or less pronounced clinical symptoms. Patients with CCDC39/40 defects, which is associated with ciliary ultrastructural microtubular disorganisation combined with inner dynein arm deficiency, tend to have poorer lung function outcomes and more severe chest CT abnormalities (14-15,21). Conversely, patients with DNAH11 defects, which are associated with normal ciliary ultrastructure (22), may have milder respiratory symptoms (14). Even though ciliary ultrastructure by TEM is normal in DNAH11, the ciliary motility pattern is markedly compromised in these patients, as ciliary activity on HSVM is characterized by stiff hyperkinetic and minimal movement (4,16). In this study, we did not find any indications that PRMC differed in any way between groups of genotypes where PCD expectedly would be related to either more severe or more mild disease. In relation to this, PRMC results has also not been shown to be associated with lung function (FEV_1_), as previously investigated by Vali and colleagues (9).

One case was an exception. This patient was the only one to show normal LR1 values and measurable, although low, TBV and had a homozygous p.His154Pro CCDC103 defect, subtle ciliary motility pattern abnormalities, a normal nNO and only a moderate ODA deficiency. This particular p.His154Pro CCDC103 defect has previously been associated with reduced-to-normal ciliary beating as well as normal range nNO (23). Even though it was only a single case and interpretation of the result obviously should be taken with caution, this finding illustrates that the PRMC method may have the ability to reflect even smaller variations in ciliary beating within PCD, which potentially could extrapolate to detectable improvement in PRMC as an outcome if ciliary function could be restored in pwPCD.

### PRMC cough clearance

This study provides novel insights into the impact of cough on pulmonary clearance in pwPCD. We found that cough clearance plays a crucial role in PRMC measurements, as coughing during the test can lead to a significant increase in PRMC values and subsequent false near-normal results. Cough clearance thereby showed to matter significantly as a pitfall for PRMC measurement. Therefore, it is important to take measures to minimize the risk of involuntary coughing during PRMC measurements and to carefully monitor cough during PRMC measurement as failure to control coughing may render the PRMC inconclusive or false negative for diagnostic purposes, and lead to false positive results with respect to any drugs expected to improve mucociliary clearance.

We found accumulated cough to result in increasing clearance towards a more normal level. These results therefore also suggest that, in addition to the obligatory, spontaneous, and unavoidable cough, pwPCD can potentially significantly optimize airway clearance through daily controlled coughing.

### Strengths and limitations

#### Strengths of the study and of the PRMC method to deliver a clinical trial outcome parameter

Our study provides new insight into the use of the PRMC test as a potential supplier of outcome parameters for clinical trials. It is well established that PRMC discriminates significantly between PCD and health (5, 7-9), and without losing diagnostic capability even if the PRMC test is shortened to a 60-minute test (9). PRMC test results related to PCD genotypes has not been described until now. We here demonstrated that PRMC is absent across various genotypes in PCD. Although evidence is weak, since only represented by one patient, we believe the study gives an indication that residual ciliary function in PCD could lead to measurable PRMC, which could be an argument for PRMC testing as provider of useful outcome parameter to detect improvement in medically enhanced ciliary function.

Additionally, PRMC can be performed relatively quickly and efficiently, with a full measurement taking only two hours from doorstep to doorstep. This ease of use makes it possible to include patients from a large hospital service area, and since even children as young as five years old can cooperate to PRMC measurements PRMC testing might also be useful in studies including younger children.

#### Limitations of the study and of the PRMC method

This study was performed using retrospective diagnostic data from our local registry collected during a 24-year period. Despite this, the sample size was relatively small since many cases were diagnosed exclusively by HSVM, TEM and nNO, and more recently with additional genetic work-up, without a supplemental PRMC test.

However, performing PRMC targeted for distinct PCD genotypes and patients with residual ciliary function could add important knowledge on how to include PRMC as an outcome parameter for future clinical trials in PCD.

Concerning the use of PRMC in clinical trials there are several limitations to consider, including radiation exposure, age restrictions, and the potential impact on peripheral deposition.

Although the radiation exposure associated with a PRMC measurement is low (<1 mSv), it is important to consider, especially if multiple measurements are planned, e.g., before, during, and at the end of a clinical trial. Yet, even with three consecutive studies performed in a patient the radiation exposure is lower than the yearly background radiation in Denmark of 4 mSv.

PRMC requires close cooperation of the patients during the controlled inhalation of the radioaerosol, and the patient must lie quiet for at least 20 minutes during the scintigraphy, which makes PRMC unsuitable for children under five years of age (5), unless a light sedation is employed. Age has also been shown to affect PRMC lung retention in healthy individuals showing children to have a faster mucociliary clearance. Currently, normal references values for PRMC lung retention do not exist for children or for TBV for separate age groups (7). However, in a clinical trial where outcome parameters from PRMC testing would be used, e.g., baseline at inclusion, during trial, and at end of trial, the patients would be their own controls, which would minimize the impact of differences associated with age.

We demonstrated a significant impact of peripheral radioaerosol deposition on cough clearance. Previously, we have shown that it also has a significant impact on lung retention values (10). Hence, minimizing peripheral deposition should be attempted.

In studies where PRMC outcome parameters are to be used, the same PI should be aimed for both at baseline, during test and at end-of-trial measurements. The deposition of the inhaled radioaerosol largely depends on aerosol particle size, inhalation pattern and the degree of airway obstruction. Since the aerosol particle size is constant and the inhalation pattern can be controlled, it is usually the degree of airway obstruction that determines the final aerosol deposition and hence the PI.

Inhomogeneous and variable peripheral obstruction is a common finding in pwPCD and measures towards reducing bronchial obstruction e.g., by considering adding beta-2 agonist inhalation to a trial may be a way to reach a similar radioaerosol deposition during a study. This addition may also decrease tendency to cough.

## CONCLUSION

With this study we evaluated more than twenty years of experience in performing diagnostic PRMC testing in Danish pwPCD with a view to use parameters from the PRMC method as possible outcomes for future clinical PCD trials.

We found that completely absent mucociliary clearance could be expected in most PCD patients. However, there is certainly potential for PRMC parameters to improve if ciliary function is partially restored, as indicated by the results shown in a single patient in the study having some ciliary function and normal PRMC lung retention and measurable, although low TBV. Involuntary cough and peripheral radioaerosol deposition were the main methodological challenges, whereas, on the contrary, controlled cough could play an important role for airway clearance in pwPCD.

Overall, the research presented highlights the potential of PRMC studies for use in future clinical trials focusing on restoration of ciliary activity and mucociliary clearance.

## Supporting information

Supplemental files: Legends for OLS movie 1a and 1b. In addition: Figure 2

OLS movie 1a) PRMC TBV in a non-PCD referral

OLS movie 1b) PRMC TBV in a patient with PCD

## Data Availability

All data produced in the present study are available upon reasonable request to the authors

## ACKNOWLEDGEMENTS

Many thanks to the staff at the Department of Clinical Physiology and Nuclear Medicine at Rigshospitalet for their skillful help in performing the pulmonary radioaerosol mucociliary studies throughout the many years.

