## Supplemental files: Legends for OLS movie 1a and 1b. In addition: Figure 2 for "Pulmonary Radioaerosol Mucociliary Clearance Parameters as Potential Outcomes in Primary Ciliary Dyskinesia Trials"

**Legend for Supplemental files: movie 1a and 1b:**

**OLS movie 1a): PRMC tracheobronchial transport in a 20 to 25 year-old non-PCD referral where PCD was later ruled out.**
Centrally deposited radioaerosol is seen to form tracer boli that are ascending from the main bronchi and further up the trachea. The tracheobronchial velocity in this individual was 6.0 mm/min.

**OLS movie 1b): PRMC tracheobronchial transport in a 6 to 11 year-old patient with PCD who has biallelic DNAH11 defect.**
The radioaerosol is centrally deposited and still, there no detectable mucociliary clearance. The tracheobronchial velocity in this patient was 0 mm/min.

**Supplemental files Figure 2: Distribution of PCD genes among the included patients with relation to their TEM and HSVM findings**

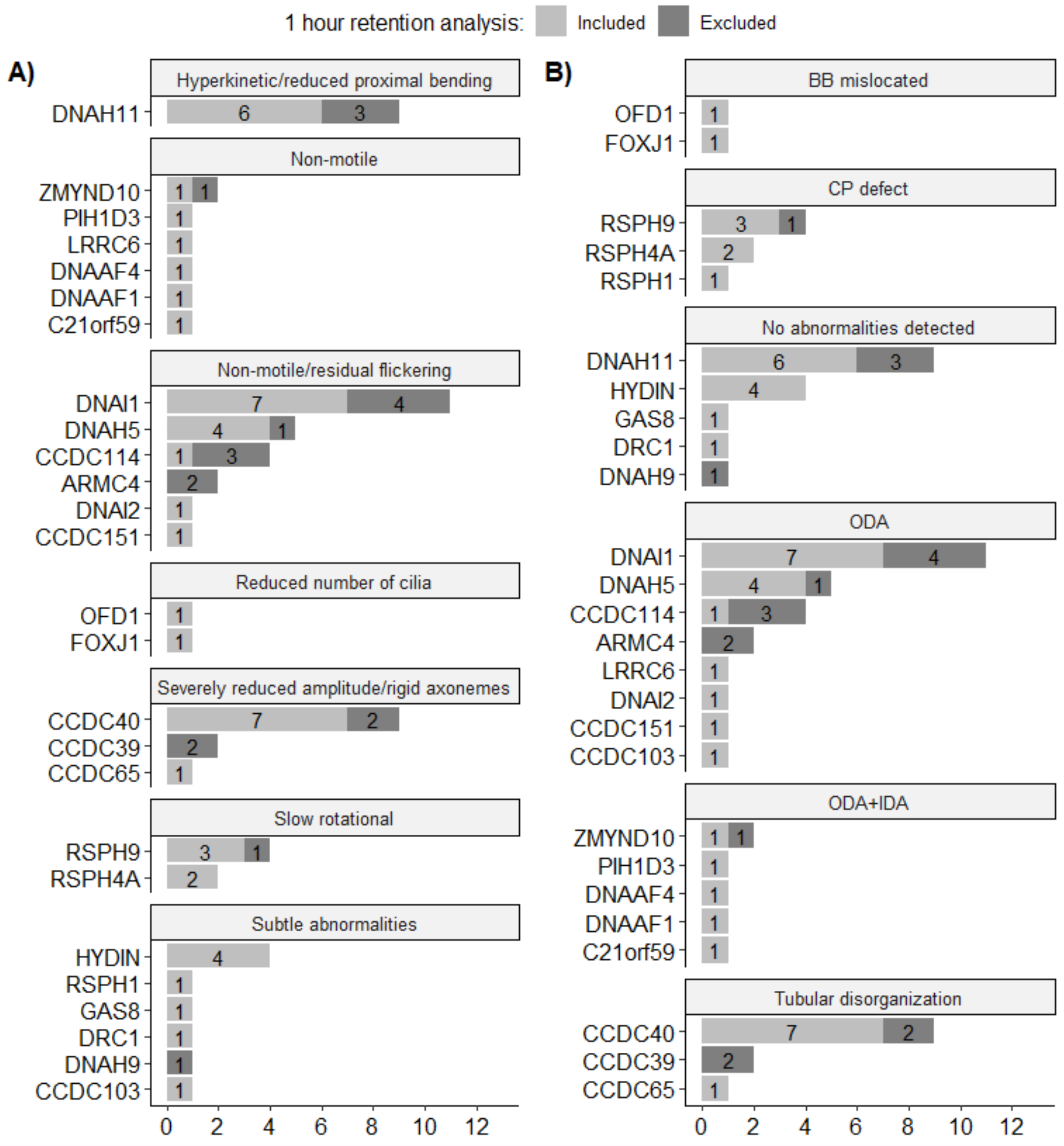


**Legend:** Distribution of genes in relation to A) motility pattern and B) ultrastructure for the included patients in the study. CCDC39 and CCDC40 defects also include IDA defect. Dark grey annotates subjects excluded in the analysis of 1 hour retention.
