## Appendix 1 for "Pulmonary Radioaerosol Mucociliary Clearance Parameters as Potential Outcomes in Primary Ciliary Dyskinesia Trials"

Procedure for calculation of reference values for PRMC and Residual Standard Deviations (RSD) based on a multiple regression model [10]:

$LR = k_0 + (k_1 \text{ x age}) + (k_2 \text{ x sex}) + (k_3 \text{ x Penetration index}) \pm SD$

Predicted LR1 (%) = 72.63 + (-0.06 x age) + (-1.72 x sex) + (25.94 x Penetration index) ± 9.01

$k_0 = 72.63$; $k_1 = -0.06$; $k_2 = -1.72$ (sex: 1 = male, 2 = female); $k_3 = 25.94$; $SD_{LR1} = 9.01$

Example:
Predicted LR1 calculated for a 6.9-year-old female (sex: 2, Penetration index: 0.46):

Predicted LR1 (%) = 72.63 + (-0.06 x 6.9) + (-1.72 x 2) + (25.94 x 0.46) ± 9.01 = 80.7 ± 9.01

**Normal range** for LR1 of this female (comprising 95% of the population):

LR1 (%) = 80.7 ± ($2.0^a$ x 9.01) i.e., between 62.7% and 98.7%,

or more correctly since abnormal mucociliary clearance can only be too slow:

LR1 (%) = 80.7 + ($1.67^b$ x 9.01) = 95.8% i.e., below 95.8% is normal.

$^a$t (N-2, P< 0.025); $^b$t (N-2, P< 0.05)

In this patient with PCD the measured lung retention after 1 hour was 100 % (i.e., nothing cleared within 1 hour)

**Calculation of RSD for LR1 in this patient**:

$RSD_{LR1} = (LR1_{measured} - LR1_{predicted}) / LR1_{SD}$

$RSD_{LR1} = (100 - 80.7) / 9.01 = +2.1$

Thus, the measured 1-hour retention lies 2.3 RSD above the predicted value, which is abnormal (i.e., all RSD above 1.67 are abnormal)
